# The Effect of Novel Exogenous Ketone Supplements on Blood Beta-Hydroxybutyrate and Glucose

**DOI:** 10.1101/2022.08.11.22278668

**Authors:** Kaja Falkenhain, Ali Daraei, Jonathan P Little

## Abstract

**Background:** Recently developed exogenous ketone monoester supplements can acutely raise blood β-OHB and lower blood glucose without the need for other nutritional modifications or invasive procedures. However, unpleasant taste and the potential for gastrointestinal discomfort may make adherence to a supplement regimen challenging. Two novel ketone supplements have been developed that promise an improved consumer experience but differ in their chemical properties; it is currently unknown how these affect blood β-OHB and blood glucose compared to the original ketone monoester supplement.

**Methods:** In a double-blind randomized cross-over pilot study, N = 12 healthy individuals (29 ± 5 years, BMI = 25 ± 4 kg/m^2^, 42% female) participated in three experimental trials with a different ketone supplement providing 10 grams of active ingredient in each trial; (i) the monoester (R)-3-hydroxybutyl (R)-3-hydroxybutyrate, (ii) D-β-hydroxybutyric acid with R-1,3-butanediol, and (iii) R-1,3-butanediol. Blood β-OHB and glucose were measured via finger prick capillary blood samples at baseline and across 240 minutes post-supplementation. Supplement acceptability, hunger, and gastrointestinal distress were assessed via questionnaires.

**Results:** β-OHB was elevated compared to baseline in all conditions. Total and incremental area under the curve (both *p* < 0.05) and peak β-OHB (*p* < 0.001) differed between conditions with the highest values seen in the ketone monoester condition. Blood glucose concentration was reduced after consumption of each supplement, with no differences in total and incremental area under the curve across supplements. Supplement acceptability was greatest for D-β-hydroxybutyric acid with R-1,3-butanediol, with no effect on hunger or evidence of gastrointestinal distress across all supplements.

**Conclusions:** Despite differences in composition, all ketone supplements tested raised β-OHB with the highest values seen after ketone monoester ingestion. Blood glucose was lowered to a similar extent across the assessed time frame with all three supplements.

## Background

Acute increases in blood beta-hydroxybutyrate (β-OHB), one of the ketone bodies produced by the liver during periods of starvation or carbohydrate restriction, can inhibit lipolysis and reduce blood glucose (1-3). However, any potential therapeutic application has traditionally been limited due to the need for ketone infusion or severe dietary restrictions to achieve nutritional ketosis. With the recent development of exogenous ketone supplements it is now possible – approximately 50 years after β-OHB’s glucose-lowering effects were first described – to acutely raise blood β-OHB concentration without the need for other nutritional modifications or invasive procedures.

Indeed, acute ingestion of oral ketone supplements potently increases blood β-OHB and immediately lowers blood glucose (4,5). In support of these findings, a recent systematic review and meta-analysis evaluating the effects of ketone salts and β-OHB-containing monoester supplements found blood glucose to be lowered across a range of study settings (e.g., during exercise, during a nutritional challenge) and populations (e.g., in healthy individuals or in individuals with prediabetes) (6). These findings suggest exogenous ketones to be a promising therapeutic strategy aimed at lowering blood glucose and improving blood glucose control in individuals with impaired glucose metabolism, particularly individuals with prediabetes or type 2 diabetes (7).

The first commercially available exogenous ketone supplements were ketone salts and ketone monoesters (5). Ketone salts are ketone molecules bound to a mineral and are typically (though not exclusively) available in powder form to be mixed with water prior to ingestion. In ketone monoester supplements, ketone molecules are bonded to a precursor (e.g., R-1,3-butanediol) and usually (although again not necessarily) available in liquid form. While both forms of exogenous ketones are utilized extensively for research purposes, they do present with potential side effects and concerns that can make adhering to a longer-term supplement regimen challenging. Ketone salts, in particular, lead to a high occurrence of gastrointestinal symptoms (6), while currently available ketone monoesters have a bitter and unpleasant taste.

Since these first iterations of exogenous ketones, novel ketone supplements have been invented that are now commercially available to the general public and promise an improved consumer experience. These supplements differ slightly in their chemical properties and/or added ingredients (e.g., flavouring) but claim efficacy for potently raising blood β-OHB. Because an improved taste and avoidance of gastrointestinal distress is of major interest if the therapeutic potential of exogenous ketones is to be explored, the aim of this study was to evaluate the response of blood β-OHB and blood glucose following acute ingestion of two novel ketone supplements (“Ketone-IQ”, R-1,3-butanediol; “Kenetik”, free D-β-hydroxybutyric acid with R-1,3-butanediol) compared to a version of the original ketone monoester (“KE4”, (R)-3-hydroxybutyl (R)-3-hydroxybutyrate), while also exploring gastrointestinal discomfort and overall supplement acceptability.

## Materials and methods

### Study overview

This was a double-blind randomized cross-over pilot study conducted in N = 12 healthy adults. The study was approved by the clinical ethics board of the University of British Columbia (H22-00083) and preregistered (NCT05273411). Interested participants had to be over the age of 18 and able to fast overnight. Exclusion criteria included being a competitive endurance athlete; following a ketogenic or low-calorie diet, a periodic fasting regimen, or regularly consuming ketogenic supplements; being unable to travel to and from the university; being unable to follow diet instructions; being pregnant or planning to become pregnant during the study; having been diagnosed with a chronic disorder of glucose or fat metabolism, including type 2 diabetes, chronic pancreatitis, or gallbladder disease; and being unable to read or communicate in English.

All participants provided written informed consent before the start of data collection. Following screening and informed consent, eligible participants participated in three experimental trials of ketone supplementation with the order of treatment randomized, balanced, and separated by at least 48 hours. The randomization schedule was created in R (using the package “randomizeBE”) with block sizes of 2 and 4, and the randomization sequence listed as “A-B-C” or a permutation thereof. The randomization sequence was revealed to the research lead and participant only on the day of testing, and the identity of the ketone supplements was blinded to both researchers and participants (see more details below).

Pre-menopausal women were tested in the follicular phase (days 3-9) to minimize any potential changes in insulin sensitivity across the menstrual cycle (8). Participants were asked to refrain from structured exercise and drinking alcohol on the day before each trial. Furthermore, participants were instructed to record their dietary intake on the day prior to their first experimental visit, and to consume similar foods and quantities on the days before the subsequent experimental visits. Dietary intake was analyzed by the research team based on self-report using MyFitnessPal (9). Participants were asked to fast overnight (at least 8 hours) before each of the experimental visits. On their first visit to the laboratory, baseline anthropometric characteristics were measured using a standard measurement tape and body composition was assessed by bio-electrical impedance analysis (TBF-410 Body Composition Analyzer, Tanita).

Upon arrival, a baseline fasting blood sample was obtained via finger prick to measure blood glucose and blood β-OHB with a handheld monitor. For 24 of the 36 study visits, capillary blood samples were analysed using the GK+ Blood Glucose and Ketone Meter (Keto Mojo). Due to supply issues related to the COVID-19 pandemic, the FreeStyle Meter (Abbott) was used for the remaining 12 study visits (see Supplementary Material for data separated by meter). The same meter was used within each participants’ visit. Blood pressure and heart rate were measured using an automated blood pressure monitoring system (OMRON 10 Series home blood pressure monitor, BP7450). After the baseline measures, the oral ketone supplement was consumed within 5 minutes. We did not attempt to match or mask the different supplement flavours or volumes given one aim of the study was to assess taste and tolerability. As such, supplements were provided unaltered and at different volumes (based on the manufacturers’ formulation). Participants were not aware which flavour and volume would correspond to which ketone supplement and all supplements were provided in opaque plastic bottles (labelled with “A”, “B”, or “C” by an independent person not involved with any other aspects of the study) such that neither the participant nor the research team were able to visually identify the supplement type. The code identifying the supplements (corresponding to “A”, “B”, and “C”) was kept in a sealed envelope and only revealed after completion of statistical analyses on blinded data.

Following consumption of the ketone supplement, additional blood samples were taken via finger prick at 15, 30, 60, 90, 120, 180, and 240 minutes. Blood pressure and heart rate were measured at the same time points. Furthermore, a questionnaire assessing gastrointestinal distress using visual analogues scales was administered 60 minutes after supplement consumption. After the final blood sample, questionnaires on hunger and fullness as well as on supplement acceptability were administered (see more details below).

### Questionnaires

We administered three different questionnaires; (i) a questionnaire on gastrointestinal symptoms 60 minutes after consumption of the ketone supplements; (ii) a questionnaire on hunger and fullness 240 minutes after ketone supplement consumption; and (iii) a questionnaire on supplement acceptability at the end of the testing day 240 minutes after consumption of the supplements.

The questionnaires on gastrointestinal distress and hunger and fullness were presented as visual analogue scales and asked participants to mark a vertical line through a 100 mm horizontal visual analogue line (ranging from “not present” to “worst imaginable”) to indicate their response to each question about nausea, urge to vomit, bloating, belching, and cramping (gastrointestinal symptoms); and how hungry, empty, and full participants felt as well as how much they thought they could eat (hunger and fullness). The questionnaire on supplement acceptability asked participants to indicate on a 7-point Likert scale how well they liked the supplement and its taste, how easy or difficult it would be and how much effort it would take to consume the supplement before every meal, and how (dis)satisfied they felt with and after taking the supplement.

### Exogenous ketone supplements

We tested three different ketone supplements; a version of the ketone monoester (R)-3-hydroxybutyl (R)-3-hydroxybutyrate (“KE4”, KetoneAid), as well as two newly developed supplements; free D-β-hydroxybutyric acid with R-1,3-butanediol in a 1:1 ratio (“Kenetik”, VitaNav) and R-1,3-butanediol (“Ketone-IQ”, HVMN). Each ketone supplement was provided at a dose of 10 g active ingredient (i.e., 10 g of (R)-3-hydroxybutyl (R)-3-hydroxybutyrate, D-β-hydroxybutyric acid with R-1,3-butanediol, or R-1,3-butanediol). This corresponded to the recommended serving size for the two new ketone supplements to be tested (D-β-hydroxybutyric acid with R-1,3-butanediol and R-1,3-butanediol) and is in line with previous pharmacokinetic studies that showed a dose of ∼ 10 g (R)-3-hydroxybutyl (R)-3-hydroxybutyrate monoester to successfully raise β-OHB within 30 minutes (5). A 10 g dose of the ketone monoester, (R)-3-hydroxybutyl (R)-3-hydroxybutyrate is equivalent to 20 mL of the “KE4” drink, which is sweetened with organic stevia and allulose and additionally contains water, citric acid, natural flavours, and potassium sorbate. A 10 g dose of D-β-hydroxybutyric acid with R-1,3-butanediol is equivalent to 237 ml of the “Kenetik” drink, which is a carbonated beverage flavoured with allulose, stevia, and natural flavours as well as water, potassium and sodium bicarbonate, and potassium sorbate as additional ingredients. A 10 g dose of R-1,3-butanediol is equivalent to 35 ml of the “Ketone-IQ” drink, which contains monk fruit extract and rebaudioside M as sweeteners alongside water, natural flavour, citric acid, potassium sorbate, and potassium benzoate as additional ingredients (see Supplementary Table 1).

### Data analysis

The sample size for this study was calculated based on prior literature on the effects of ketone supplements; a previous pharmacokinetic study exploring the effects of a (R)-3-hydroxybutyl (R)-3-hydroxybutyrate monoester provided at a comparable dose of ∼10 g in healthy adults found that blood β-OHB was raised to 1.4 ± 0.6 mM within 30 minutes, and to an average of 0.5 ± 0.5 mM across the entire 4-hour post-supplementation period (5), which represents an effect size of *d* > 1.0 compared to baseline. Using the outcome data from this study (N = 15) and assuming 80% power with an alpha level of 0.05 and a moderate correlation between repeated measures of 0.5, a total sample size of N = 6 would be sufficient to detect an increase in average blood β-OHB compared to baseline (calculated using G*Power v3.1). In order to allow for a more nuanced evaluation of differences between ketone supplements and to account for any dropouts or missing blood samples, we enrolled 12 participants (aiming for equal males and females), which is in line with the sample size of previous pharmacokinetic studies of ketone supplements (5,10).

The primary outcome for this study was total blood β-OHB as represented by the area under the curve across 4 hours. Secondary outcomes included blood glucose, blood pressure, heart rate, taste and supplement acceptability, hunger and fullness, and gastrointestinal tolerability; all measured across the 240-minute post-supplementation period.

Data were analyzed in R (11). Statistical significance was set at *p* < 0.05. There were no dropouts and only two participants had missing values for a subset of blood pressure measurements on one of the testing visits due to equipment malfunction; no data were imputed as per contemporary guidelines (12). The (incremental) area under the curve was calculated using the composite trapezoid rule and subtracting areas with values opposite to the direction of change (e.g., β-OHB concentration below baseline when calculating incremental area under the curve). Differences in blood β-OHB and blood glucose across testing conditions and time were determined using a linear mixed model with condition, time, the interaction thereof, and the visit order included as fixed factors, the baseline value as a covariate, and a random intercept for participant. Area under the curve, incremental area under the curve, time to reach peak blood β-OHB as well as peak blood β-OHB were compared using a linear mixed model with condition and the visit order included as fixed factors, a random intercept for participant, and a planned contrast between the ketone monoester and each of the other supplements in case of overall statistical significance. Due to non-normally distributed data derived from the supplement acceptability questionnaire, these data were analyzed using the non-parametric Kruskal-Wallis test with post-hoc pairwise Wilcoxon rank sum tests between the ketone monoester and each of the other two supplements and Bonferroni correction for multiple testing in case of overall statistical significance. Finally, the linear relationship between blood β-OHB and blood glucose was assessed using a Pearson correlation. All raw data (mean ± SD for all measures and timepoints) are presented in the Supplementary Material.

## Results

### Participants

Participant baseline characteristics are shown in Table 1. All participants completed all conditions. Dietary intake did not differ between days prior to testing across supplements (see Supplementary Table 2).

**Table 1.** Participant baseline characteristics.

|  | Total | Male | Female |
| --- | --- | --- | --- |
| N (%) | 12 (100) | 7 (58) | 5 (42) |
| Age, mean (SD), years | 29.1 (4.9) | 28.1 (3.2) | 30.4 (6.9) |
| Weight, mean (SD), kg | 71.6 (13.8) | 79.1 (12.6) | 61.2 (7.3) |
| BMI, mean (SD), kg/m<sup>2</sup> | 24.5 (4.4) | 25.6 (5.0) | 23.0 (3.3) |
| Body fat, mean (SD), % | 19.6 (9.0) | 15.7 (9.3) | 25.1 (5.6) |
| Fat mass, mean (SD), kg | 14.3 (8.5) | 13.3 (10.4) | 15.7 (5.5) |
| Lean body mass, mean (SD), kg | 57.3 (11.2) | 65.7 (5.5) | 45.5 (2.3) |

### Blood beta-hydroxybutyrate

As expected, blood β-OHB was elevated above baseline in all conditions following consumption of the supplement (see Supplementary Table 3). There were differences in β-OHB over time between supplements with a statistically significant interaction effect (*p* < 0.001) (Figure 1A).

**Figure 1.**
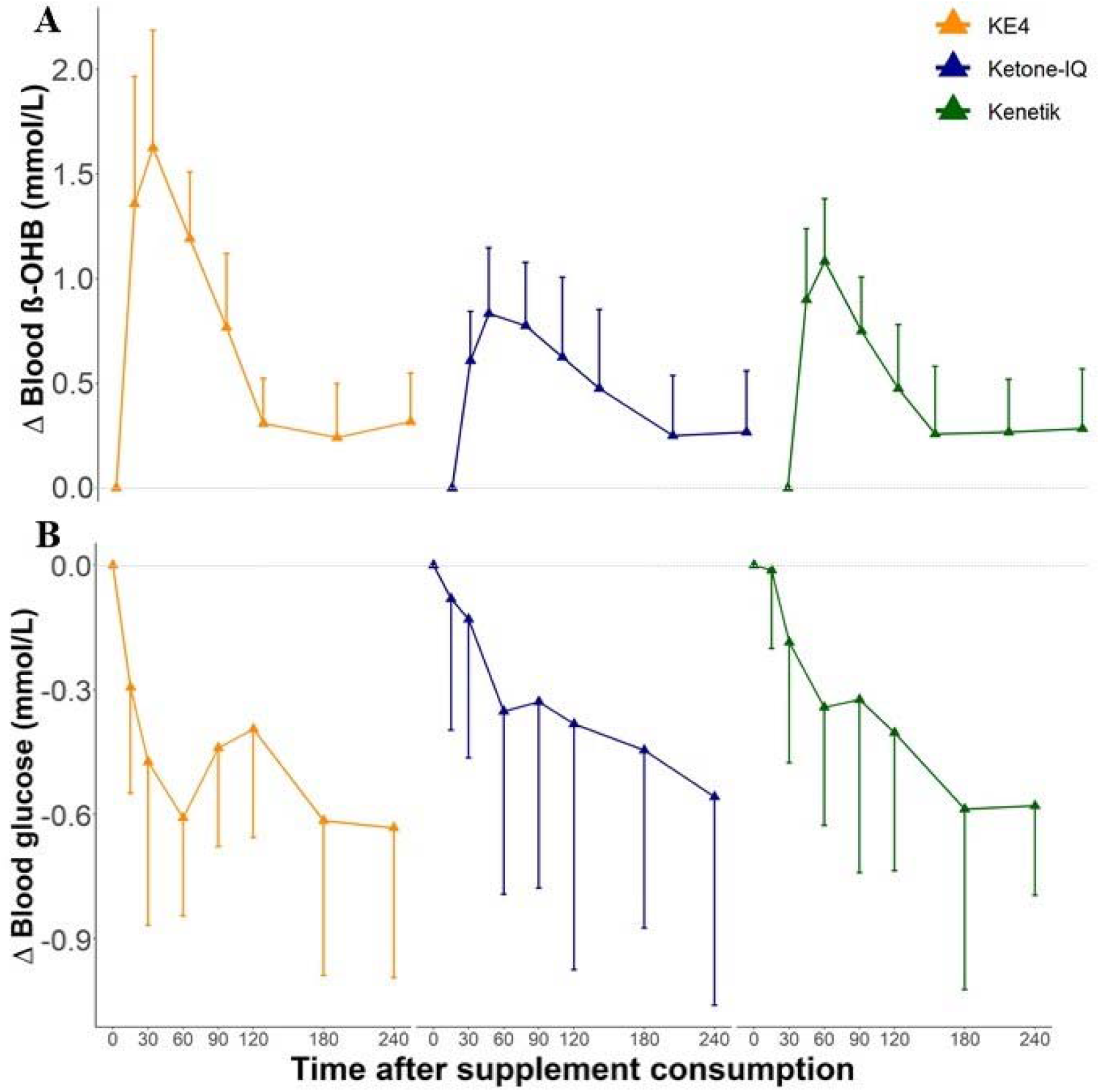
Responses of (A) blood beta-hydroxybutyrate (β-OHB) and (B) glucose concentration following consumption of three different ketone supplements (“KE4” left/orange; “Ketone-IQ”, center/blue; “Kenetik”, right/green) across 240 minutes. Supplements were consumed in the fasted state; β-OHB and glucose were assessed via finger prick capillary blood samples. Data presented as mean ± SD representing change from baseline (N = 12). See Supplementary Figures 1 & 2 for individual data.

There was a statistically significant difference in total β-OHB area under the curve (F_(2,20)_ = 4.3892, *p* = 0.03) across supplements, with “KE4” having a greater area under the curve (217 ± 51 mmol/L x 240 min) than “Ketone-IQ” (−41 mmol/L x 240 min, 95% CI: -67 to -15 mmol/L x 240 min, *p* = 0.01) but not when compared to “Kenetik” (−25 mmol/L x 240 min, 95% CI: -51 to 1 mmol/L x 240 min, *p* = 0.09). Similarly, there was a statistically significant difference in incremental β-OHB area under the curve (F_(2,20)_ = 3.5479, *p* = 0.05), which was higher after “KE4” (161 ± 51 mmol/L x 240 min) compared to both “Ketone-IQ” (−43 mmol/L x 240 min, 95% CI: - 80 to -7 mmol/L x 240 min, *p* = 0.04) and “Kenetik” (−47 mmol/L x 240 min, 95% CI: - 84 to -11 mmol/L x 240 min, *p* = 0.03). In contrast, there was no statistically significant difference in the time to reach peak β-OHB values across conditions (*p* = 0.27). There was, however, a significant difference in absolute peak β-OHB concentration (F_(2,20)_ = 13.3848, *p* < 0.001) between “KE4” (2.0 ± 0.6 mmol/L) and each of the other two novel supplements, “Ketone-IQ” (−0.8 mmol/L, 95% CI: -1.1 to -0.5 mmol/L, *p* < 0.001) and “Kenetik” (−0.5 mmol/L, 95% CI: -0.8 to -0.2 mmol/L, *p* < 0.01) (Figure 2).

**Figure 2.**
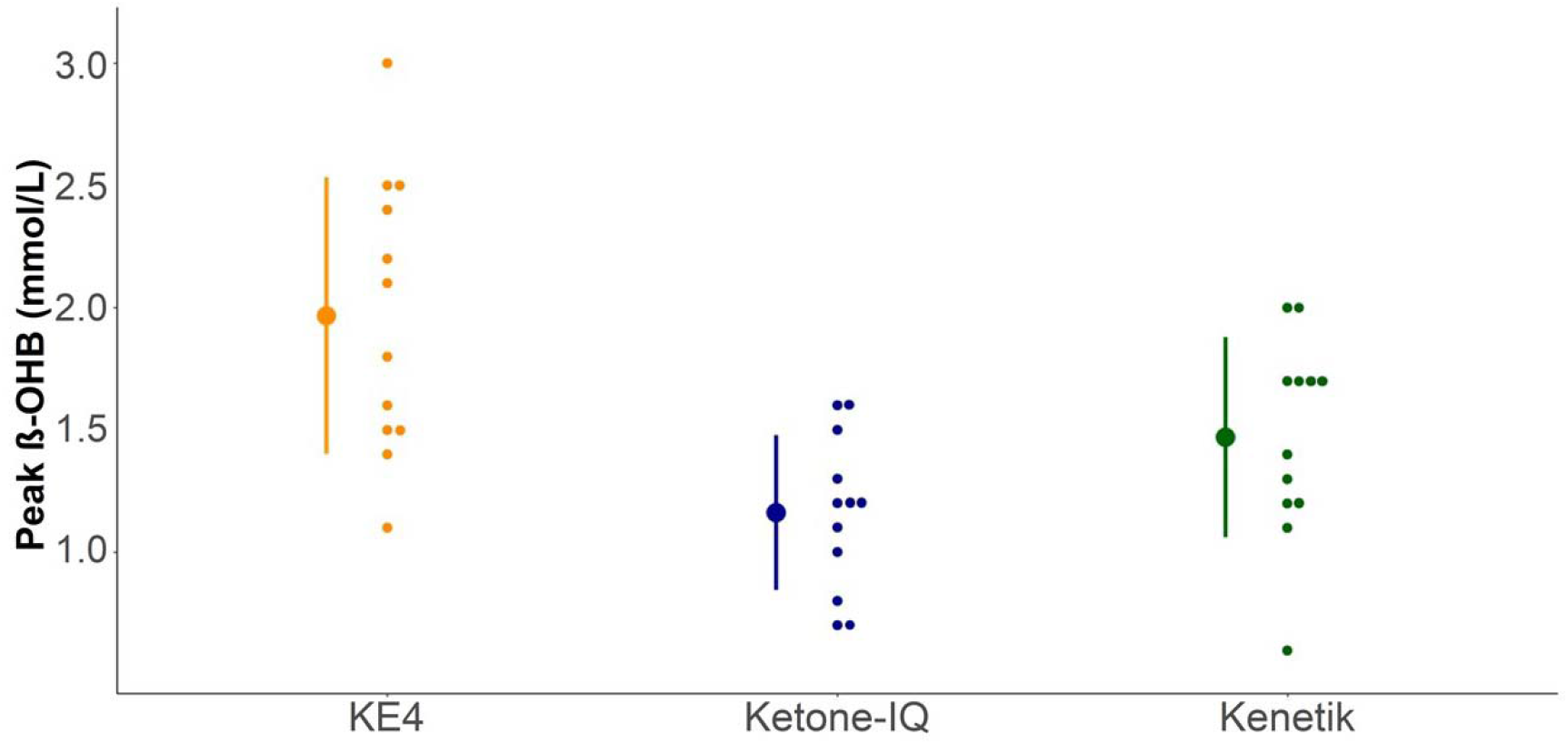
Peak beta-hydroxybutyrate (β-OHB) concentrations across 240 minutes after consumption of three different ketone supplements (“KE4”, left/orange; “Ketone-IQ”, center/blue; “Kenetik”, right/green). Supplements were consumed in the fasted state; β-OHB concentration was assessed via finger prick capillary blood samples. Individual data presented alongside mean and standard deviation (N = 12).

### Blood glucose

Blood glucose concentration decreased following supplement consumption across all conditions (see Supplementary Table 4). There was a statistically significant main effect of time and condition (both *p* < 0.001), but no interaction effect (*p* = 0.11) (Figure 1B). Neither the area under the curve (*p* = 0.56) nor the incremental area under the curve (*p* = 0.26) (Figure 3) differed between supplements.

**Figure 3.**
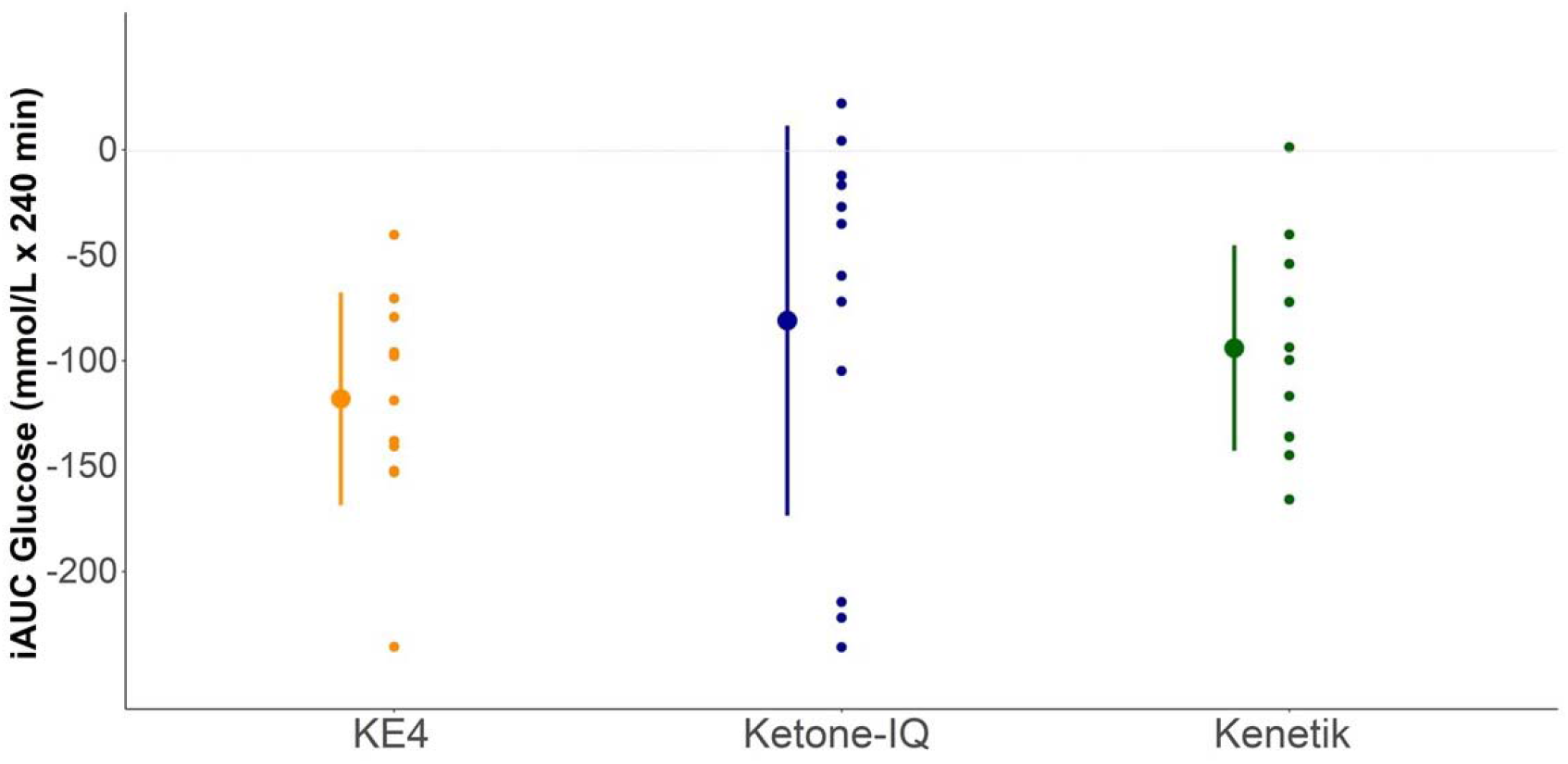
Glucose incremental area under the curve across 240 minutes after consumption of three different ketone supplements (“KE4”, left/orange; “Ketone-IQ”, center/blue; “Kenetik”, right/green). Supplements were consumed in the fasted state; glucose was assessed via finger prick capillary blood samples. Individual data presented alongside mean and standard deviation (N = 12).

### Blood pressure and heart rate

There was neither an interaction effect nor a main effect of time or condition in (diastolic or systolic) blood pressure (see Supplementary Table 5). There was a statistically significant interaction effect for heart rate (*p* = 0.04) (see Supplementary Table 6); however, testing of specific contrasts revealed no statistically significant differences between supplements at any timepoint.

### Questionnaire data

Gastrointestinal distress was low across all conditions with a median value of 0.0 across all measures (e.g., bloating, urge to vomit) and supplements except nausea which showed a median value of 0.4 (on a scale of 0 to 10) after consumption of “KE4” (see Supplementary Table 7). Due to the low incidence of gastrointestinal symptoms and the highly skewed data distribution, we did not perform statistical significance testing on these outcomes. Measures of hunger and fullness were consistent across supplements and did not differ statistically (see Supplementary Table 8).

Supplement acceptability differed across supplements (Table 2). Across all measures of acceptability (e.g., taste, overall satisfaction), “Kenetik” scored significantly higher than “KE4”. In particular, measures related to the taste of the supplement and the hypothetical ease of consistently taking the supplement were perceived as significantly better following consumption of “Kenetik” compared to “KE4”. While “Ketone-IQ” scored higher than “KE4” numerically across all measures, post-hoc testing revealed that there was no statistical significance compared to “KE4” after correcting for multiple testing.

**Table 2.**
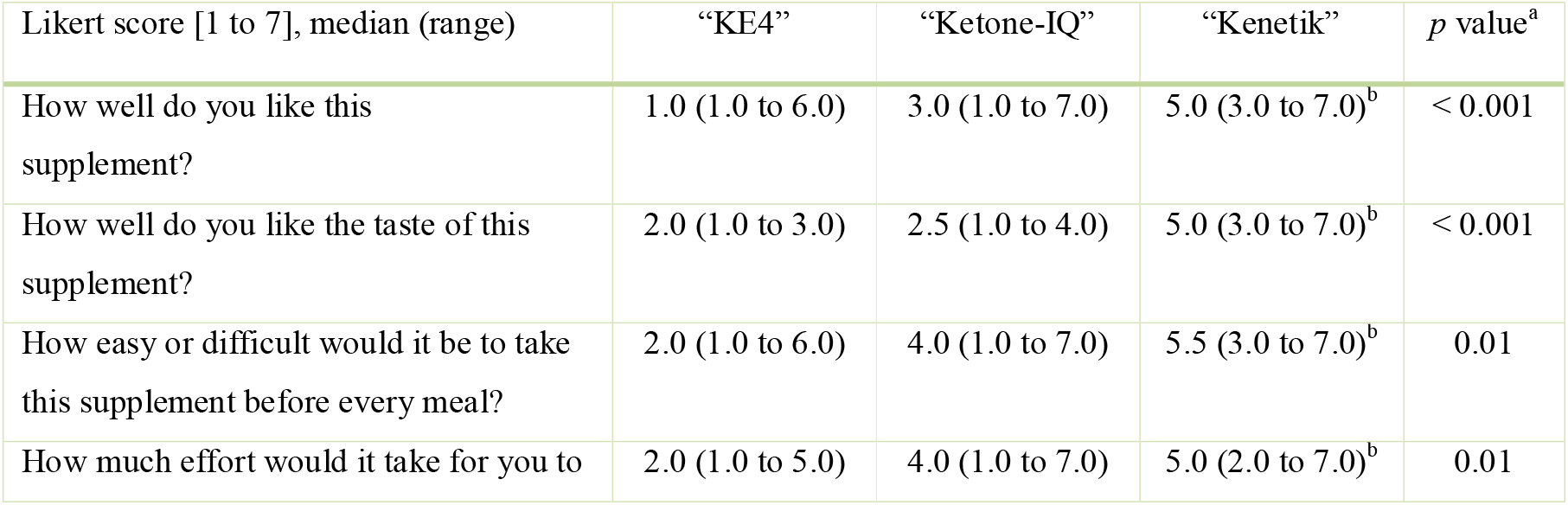
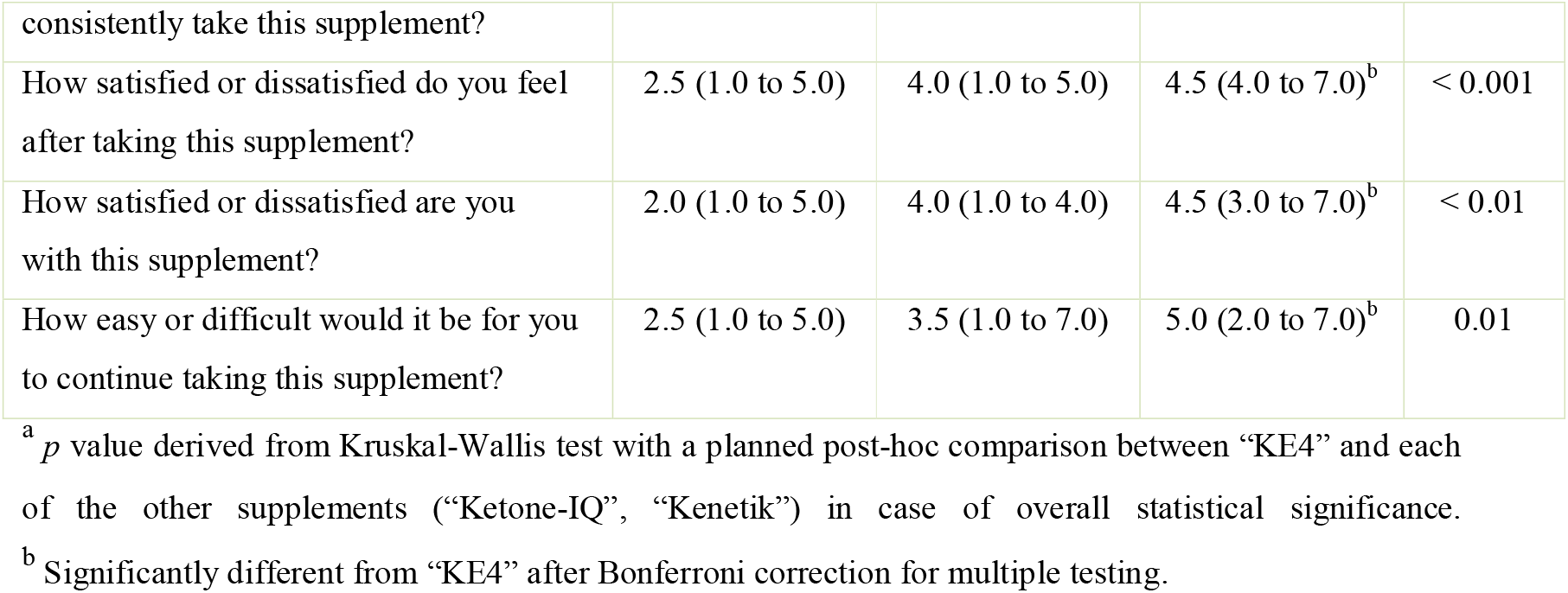
Supplement acceptability across different ketone supplements.

### Exploratory analyses

The lowest blood glucose concentrations were reached at the end of the post-supplementation period across all supplements (which can be hypothesized to be partially due to the prolonged period of remaining fasted) – however, the first glucose minimum occurred 60 minutes after supplement ingestion in all conditions (before glucose values temporarily plateaued/rose to then further decrease in a seemingly biphasic response pattern). Therefore, we explored the lowest “first minimum” blood glucose value as well the incremental area under the curve from baseline to 60 minutes across supplements (see Supplementary Table 9).

Blood glucose values 60 minutes after supplement consumption were similar across groups (*p* = 0.27); however, change from baseline to 60 minutes was near statistical significance (*p* = 0.09) with both “Ketone-IQ” (0.3 mmol/L, 95% CI: 0.01 to 0.5 mmol/L, *p* = 0.06) and “Kenetik” (0.3 mmol/L, 95% CI: 0.02 to 0.5 mmol/L, *p* = 0.05) showing a smaller initial decrease of blood glucose when compared to “KE4” (−0.6 ± 0.2 mmol/L). Similarly, the total area under the curve was comparable across supplements (*p* = 0.27), whereas there was a statistically significant difference in incremental area under the curve (*p* = 0.02), which was smaller after consumption of “Ketone-IQ” (13 mmol/L x 60 min, 95% CI: 4 to 23 mmol/L x 60 min, *p* = 0.02) and “Kenetik” (15 mmol/L x 60 min, 95% CI: 5 to 24 mmol/L x 60 min, *p* = 0.01) than after “KE4” (−24 ± 13 mmol/L x 60 min).

Finally, we evaluated whether the degree of “β-OHB exposure” (i.e., the incremental area under the curve) correlated with the overall degree of glucose-lowering observed (i.e., the incremental area under the curve). There was a statistically significant correlation between β-OHB exposure and overall glucose-lowering effect for “KE4” (r_(10)_ = -0.61, 95% CI: -0.88 to -0.06, *p* = 0.03) (Figure 4). In contrast, there were no statistically significant relationships between the two measures for “Ketone-IQ” (r_(10)_ = -0.23, 95% CI: -0.71 to 0.40, *p* = 0.47) or “Kenetik” (r_(10)_ = -0.33, 95% CI: -0.76 to 0.30, *p* = 0.29).

**Figure 4.**
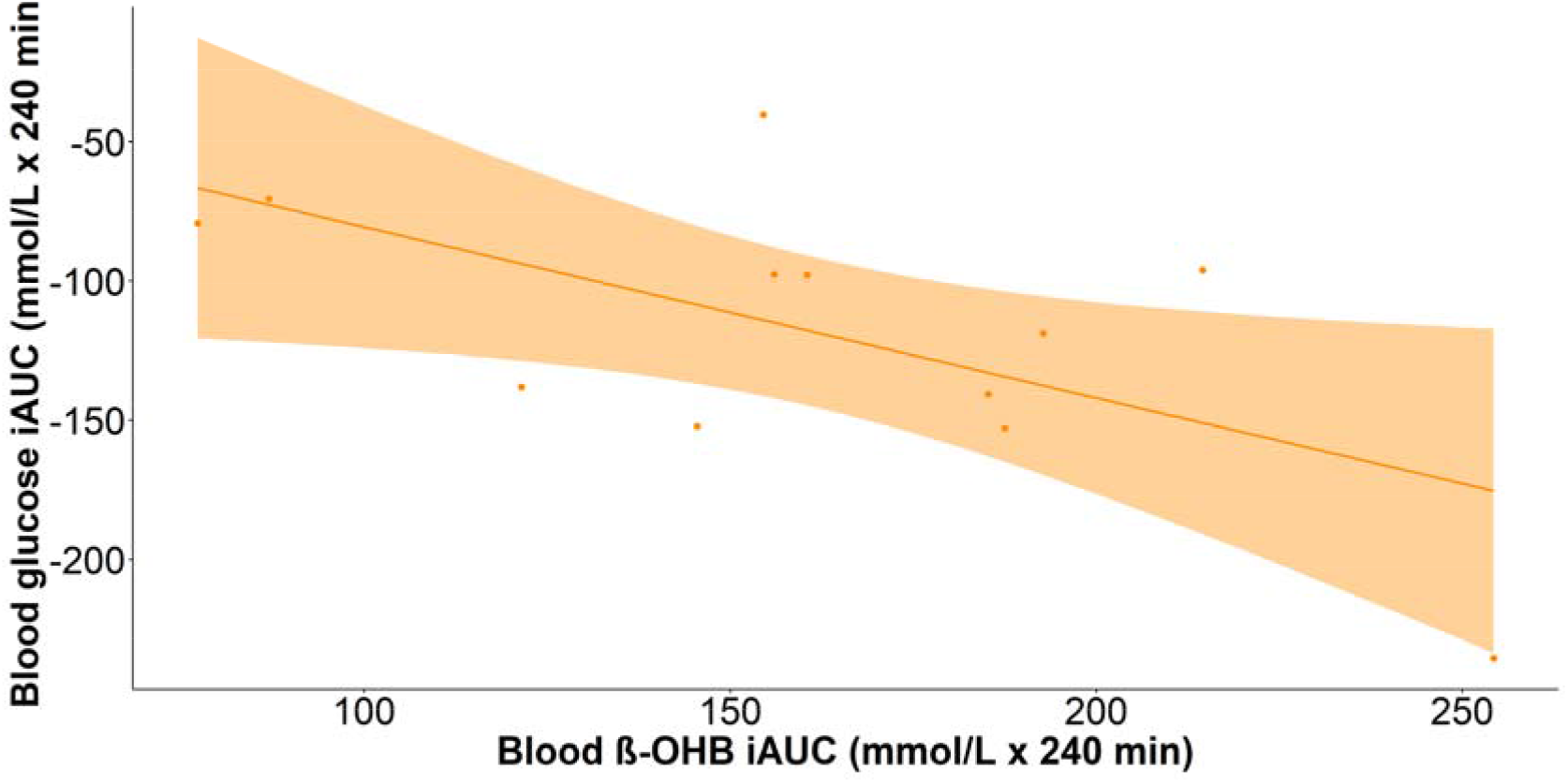
Correlation between blood beta-hydroxybutyrate (β-OHB) incremental area under the curve and blood glucose incremental area under the curve across 240 minutes following consumption of the ketone monoester “KE4”. The supplement was consumed in the fasted state; glucose and β-OHB concentrations were assessed via finger prick capillary blood samples. Individual data presented alongside the line of best fit with corresponding 95% confidence interval (N = 12).

## Discussion

In this study, we present evidence for the β-OHB-raising and glucose-lowering effect (and the time course thereof) of two novel exogenous ketone supplements (D-β-hydroxybutyric acid with R-1,3-butanediol mixture, and R-1,3-butanediol alone) compared to the well-established ketone monoester (R)-3-hydroxybutyl (R)-3-hydroxybutyrate. We also share data on gastrointestinal distress and overall supplement acceptability comparing these three ketone supplements.

As has been shown in previous studies (5,6), the ketone monoester supplement elevated blood β-OHB concentrations compared to baseline. Similarly, the novel ketone supplements raised blood β-OHB; however, to an overall lesser degree (i.e., when exploring the total or incremental area under the curve). Furthermore, the ketone monoester led to a greater first-phase increase in β-OHB and reached a higher peak β-OHB value than either of the other ketone supplements. While the physiological relevance (e.g., in terms of signalling or other downstream properties) of this steep rise and fall (compared to a slower rise and more prolonged elevation) is unknown, we did see the glucose response to ingestion of the exogenous ketones mirror the rise in β-OHB. That is, the first decrease in glucose was more pronounced after ingestion of the ketone monoester when compared to the other two supplements. In contrast, however, the overall glucose-lowering effect did not statistically differ between supplements (i.e., the total and incremental area under the curve were comparable).

Interestingly, the glucose response following consumption of the ketone supplements was closely in line with what would be predicted based on previously published data on the relation between blood β-OHB elevation and lowered blood glucose. For example, a recent systematic review and meta-analysis conducted a meta-regression across 33 published comparisons evaluating ketone salts and/or ketone monoesters consumed in a fasted state in a within-group analysis (i.e., comparing the response following ingestion of a ketone supplement to baseline) (6). In this analysis, the average decrease in blood glucose (as assessed across the immediate post-supplementation period, comparable to the 240 minutes following supplement consumption in the present study) was regressed on either average or peak blood β-OHB. Figure 5 (see also Supplementary Table 10) shows the observed *vs* predicted glucose response as observed in this study compared to the historic data as reported in the previous meta-analysis (6). The tight relation between blood β-OHB concentration and decrease in blood glucose further supports the direct glucose-lowering effect of acute elevations in blood β-OHB induced by a variety of exogenous ketone supplements, including those not directly containing β-OHB such as R-1,3-butanediol.

**Figure 5.**
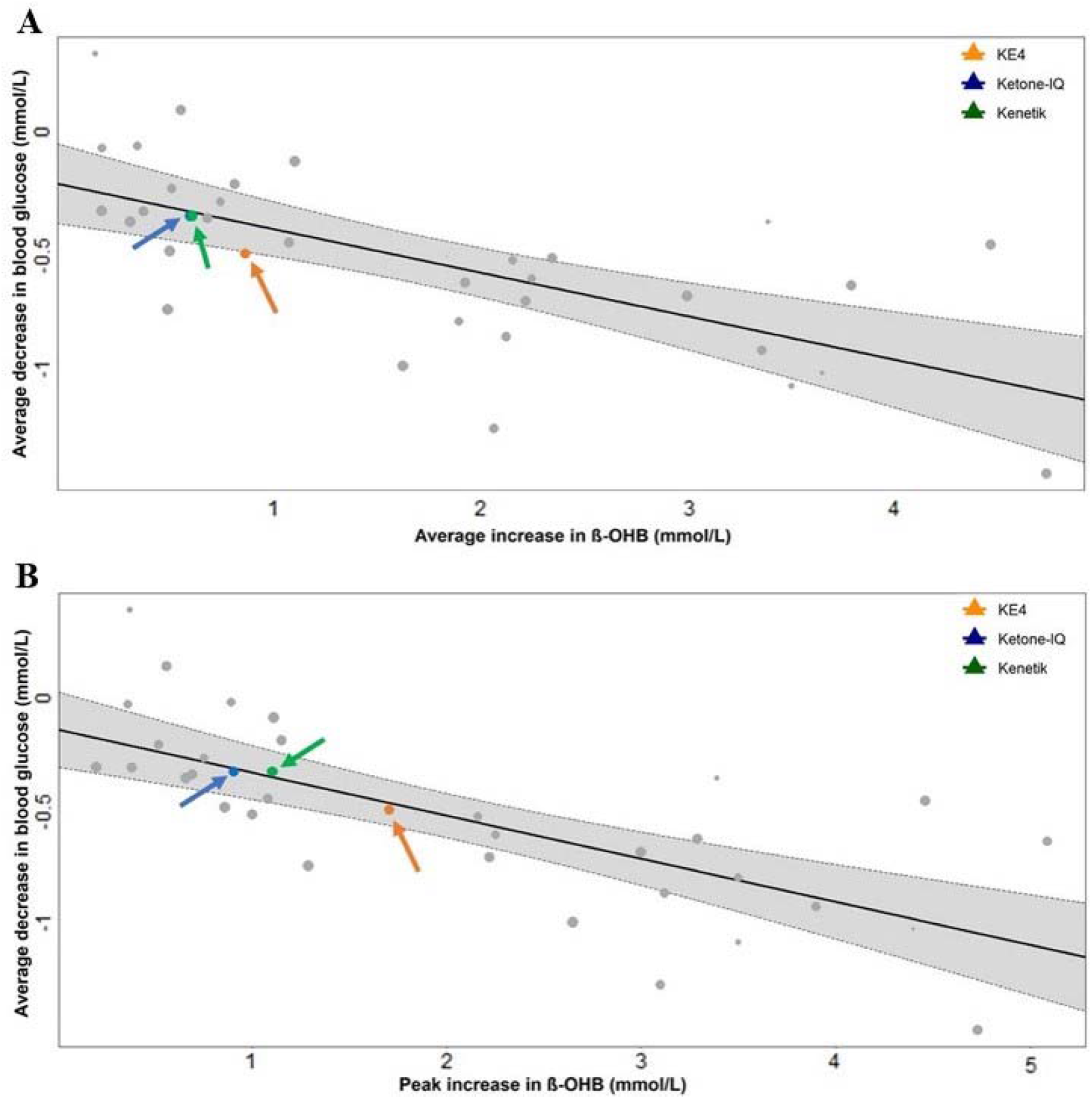
Figure redrawn from Falkenhain et al, 2022 (6). Meta-regression of (A) the average increase in blood beta-hydroxybutyrate (β-OHB) or (B) the peak increase in β-OHB as potential predictors of the average decrease in blood glucose after acute consumption of exogenous ketone (salt or monoester) supplements across 33 previous comparisons from the literature and the 3 supplements used in this study. All comparisons were conducted in a fasted stated and within-group (i.e., after ketone supplementation compared to baseline). Previously published study-level data presented alongside regression line and corresponding 95% confidence interval; results from this study have been added as coloured circles (“KE4”, orange; “Ketone-IQ”, blue; “Kenetik”, green) and are indicated by arrows.

Although some hypotheses exist, the exact mechanism by which exogenous ketones lower blood glucose has not been fully elucidated, and we did not address this in the current investigation. It has been suggested that ketones directly increase insulin secretion (1); decrease gluconeogenesis and hepatic glucose output, for example by inhibiting lipolysis (2,3,13) and reducing gluconeogenic precursors (14); improve insulin sensitivity (15-17), increase peripheral glucose uptake, and/or modulate the sympathetic nervous system (18). However, further research is needed to investigate the mechanistic underpinnings that underlie the observed glucose-lowering effect of exogenous ketones in different metabolic (e.g., fed *vs* fasted) or disease (e.g., type 2 diabetes *vs* healthy) states. Similarly, the pharmacokinetics of different ketone supplements and the physiological relevance thereof is currently unknown and warrants further research.

This study is somewhat limited by the absence of a placebo group, which was decided upon for two main reasons; (i) the primary outcome of interest being how the novel ketone supplements compare to the well-established β-OHB-raising and glucose-lowering effect of the ketone monoester; and (ii) the lack of a suitable “inert” placebo that would be an appropriate comparison across all supplements. The latter in particular was complicated given the different volumes and appetitive characteristics of the different supplements employed, as well as the glucose-affecting properties of potential comparator options (e.g., carbohydrates or protein independently affecting blood glucose, fatty acids potentially affecting production of blood ketones, nutrient-deplete water not matched for caloric content and so on). The present study therefore does not allow us to evaluate the effects of the ketone supplements on blood β-OHB and blood glucose compared to a time-matched period of fasting. Furthermore, our study population consisted of young, healthy adults in a fasted state and consequentially, the observed kinetic properties of the ketone supplements cannot be extrapolated to other populations (e.g., older adults, individuals with type 2 diabetes) and other metabolic states (e.g., postprandial, during exercise).

In conclusion, all three exogenous ketone supplements employed in this study induce exogenous ketosis and exhibit glucose-lowering effects with no distinct evidence of causing gastrointestinal distress. Supplements containing 10 grams of free D-β-hydroxybutyric acid combined with R-1,3-butanediol or 10 grams of R-1,3-butanediol alone lead to lower peak β-OHB concentrations when compared to a matched 10 g dose of the (R)-3-hydroxybutyl (R)-3-hydroxybutyrate ketone monoester, although the absolute glucose-lowering effects appeared similar. Furthermore, overall supplement acceptability was highest with the carbonated beverage drink combining D-β-hydroxybutyric acid with R-1,3-butanediol, potentially supporting its use in future research aiming to explore the feasibility of prolonged ketone supplementation in the real world.

## Data Availability

Data and corresponding code are publicly available under this link: https://osf.io/wvxf4/?view_only=2a2bc242fabd41babeba1b41d04652a5

## Authors’ contribution

Conceptualization, all authors; methodology, K.F., J.P.L.; formal analysis, K.F.; investigation, all authors; data curation, K.F., A.D.; writing—original draft preparation, K.F.; writing—review and editing, all authors; visualization, K.F.; supervision, J.P.L.; project administration, K.F., J.P.L.; funding acquisition, J.P.L. All authors have read and agreed to the published version of the manuscript

## Institutional review board statement

The study was approved by the clinical ethics board of the University of British Columbia (H22-00083) and preregistered (NCT05273411).

## Disclosures statement

J.P.L. is supported by a Michael Smith Foundation for Health Research (MSFHR) Scholar Award (16890) and a Killam Accelerator Research Fellowship. J.P.L. is volunteer Chief Scientific Officer for the not-for-profit Institute for Personalized Therapeutic Nutrition. J.P.L. holds founder shares in Metabolic Insights Inc., a for-profit company that developed non-invasive metabolic monitoring devices.

## Additional information

### Funding

This work was funded by a Canadian Institutes of Health Research (CIHR) Project Grant to J.P.L. (PJT-169116).

## Notes

### Clinical Trial

NCT05273411

## References

1. Madison LL, Mebane D, Unger RH, Lochner A. The hypoglycemic action of ketones. II. Evidence for a stimulatory feedback of ketones on the pancreatic β cells. J Clin Invest. 1964;43(3):408–15.

2. Senior B, Loridan L. Direct regulatory effect of ketones on lipolysis and on glucose concentrations in man. Nature. 1968;219(5149):83–4.

3. Taggart AKP, Kero J, Gan X, Cai TQ, Cheng K, Ippolito M, Ren N, Kaplan R, Wu K, Wu TJ, et al. (D)-β-hydroxybutyrate inhibits adipocyte lipolysis via the nicotinic acid receptor PUMA-G. J Biol Chem. 2005;280(29):26649–52.

4. O’Malley T, Myette-Cote E, Durrer C, Little JP. Nutritional ketone salts increase fat oxidation but impair high-intensity exercise performance in healthy adult males. Appl Physiol Nutr Metab. 2017;42(10):1031–5.

5. Stubbs BJ, Cox PJ, Evans RD, Santer P, Miller JJ, Faull OK, Magor-Elliott S, Hiyama S, Stirling M, Clarke K. On the metabolism of exogenous ketones in humans. Front Physiol. 2017;8:848.

6. Falkenhain K, Daraei A, Forbes SC, Little JP. Effects of exogenous ketone supplementation on blood glucose: a systematic review and meta-analysis. Adv Nutr. 2022;nmac036.

7. Walsh JJ, Myette-Côté É, Neudorf H, Little JP. Potential therapeutic effects of exogenous ketone supplementation for type 2 diabetes: a review. Curr Pharm Des. 2020;26(9):958–69.

8. Diamond MP, Simonson DC, DeFronzo RA. Menstrual cyclicity has a profound effect on glucose homeostasis. Fertility and Sterility. 1989;52(2):204–208.

9. MyFitnessPal.com [online]. Free calorie counter, diet & exercise journal. 2022. Available from: https://www.myfitnesspal.com/ [Accessed 20 June 2022].

10. Crabtree CD, Blade T, Hyde PN, Buga A, Kackley ML, Sapper TN, Panda O, Roa-Diaz S, Anthony JC, Newman JC, et al. Bis hexanoyl (R)-1,3-butanediol, a novel ketogenic ester, acutely increases circulating R- and S-ß-Hydroxybutyrate concentrations in healthy adults. J Am Nutr Assoc. 2022;3:1–9.

11. R Core Team. A language and environment for statistical computing [online]. R Foundation for Statistical Computing. 2020. Available from: https://www.r-project.org/ [Accessed 20 June 2022].

12. Chakraborty H. A mixed model approach for intent-to-treat analysis in longitudinal clinical trials with missing values [online]. 2009. Available from: http://www.rti.org/publication/mixed-model-approach-intent-treat-analysis-longitudinal-clinical-trials-missing-values

13. Reaven G. All obese individuals are not created equal: insulin resistance is the major determinant of cardiovascular disease in overweight/obese individuals. Diabetes Vasc Dis Res. 2005;2(3):105–12.

14. Soto-Mota A, Norwitz NG, Evans RD, Clarke K. Exogenous D-β-hydroxybutyrate lowers blood glucose in part by decreasing the availability of L-alanine for gluconeogenesis. Endocrinol Diabetes Metab. 2022:5(1):e00300.

15. Santomauro ATMG, Boden G, Silva MER, Rocha DM, Santos RF, Ursich MJM, Strassmann PG, Wajchenberg BL. Overnight lowering of free fatty acids with Acipimox improves insulin resistance and glucose tolerance in obese diabetic and nondiabetic subjects. Diabetes. 1999;48(9):1836–41.

16. Saloranta C, Groop L, Ekstrand A, Franssila-Kallunki A, Eriksson J, Taskinen MR. Different acute and chronic effects of Acipimox treatment on glucose and lipid metabolism in patients with type 2 diabetes. Diabet Med. 1993;10(10):950–7.

17. Worm D, Henriksen JE, Vaag A, Thye-Ronn P, Melander A, Beck-Nielsen H. Pronounced blood glucose-lowering effect of the antilipolytic drug Acipimox in noninsulin-dependent diabetes mellitus patients during a 3-day intensified treatment period. J Clin Endocrinol Metab. 1994;78(3):717–21.

18. Kimura I, Inoue D, Maeda T, Hara T, Ichimura A, Miyauchi S, Kobayashi M, Hirasawa A, Tsujimoto G. Short-chain fatty acids and ketones directly regulate sympathetic nervous system via G protein-coupled receptor 41 (GPR41). Proc Natl Acad Sci. 2011;108(19):8030–5.

