## Supplemental Tables and Figures for "The Effect of Novel Exogenous Ketone Supplements on Blood Beta-Hydroxybutyrate and Glucose"

**Supplementary Table 1. Composition of ketone supplements employed in this study**

|  | “KE4” | “Ketone-IQ” | “Kenetik” |
| --- | --- | --- | --- |
| Ketone molecule | (R)-3-hydroxybutyl (R)-3-hydroxybutyrate | R-1,3-butanediol | free R-β-hydroxybutyric acid with R-1,3-butanediol in a 1:1 ratio |
| Other ingredients | water, stevia, allulose, citric acid, natural flavours, potassium sorbate | water, monk fruit extract, rebaudioside M, natural flavour, citric acid, potassium sorbate, potassium benzoate | water, allulose, stevia, natural flavours, potassium bicarbonate, sodium bicarbonate, potassium sorbate |
| Volume (per 10 g active ingredient) | 20 mL | 35 mL | 237 mL |
| Nutritional content (per 10 g active ingredient) | 48 kcal | 70 kcal | 60 kcal |

**Supplementary Table 2. Dietary intake prior to testing days**

|  | “KE4” | “Ketone-IQ” | “Kenetik” | *p* value^a^ |
| --- | --- | --- | --- | --- |
| Energy intake, mean (SD), kcal | 1631 (466) | 1752 (597) | 1788 (792) | 0.60 |
| Carbohydrate intake |  |  |  |  |
| Mean (SD), g | 190 (64) | 207 (80) | 195 (59) | 0.76 |
| Mean (SD), % | 49 (16) | 49 (13) | 45 (15) | 0.46 |
| Fat intake |  |  |  |  |
| Mean (SD), g | 63 (33) | 67 (35) | 77 (61) | 0.43 |
| Mean (SD), % | 34 (12) | 34 (13) | 34 (14) | 0.97 |
| Protein intake |  |  |  |  |
| Mean (SD), g | 73 (36) | 79 (38) | 83 (42) | 0.44 |
| Mean (SD), % | 18 (8) | 18 (7) | 19 (5) | 0.93 |

^a^ *p* value derived from linear mixed model with condition and visit order included as fixed factors, a random intercept for participant, and a planned contrast between “KE4” and each of the other supplements (“Ketone-IQ”, “Kenetik”) in case of overall statistical significance.

**Supplementary Table 3. Beta-hydroxybutyrate response following different ketone supplements**

|  | “KE4” | “Ketone-IQ” | “Kenetik” | *p* value |
| --- | --- | --- | --- | --- |
| AUC, mean (SD), mmol/L x 240 min | 217 (51) | 176 (59)^a^ | 192 (63) | 0.03^b^ |
| iAUC, mean (SD), mmol/L x 240 min | 161 (51) | 118 (64)^a^ | 114 (51)^a^ | 0.05^b^ |
| Time to peak, mean (SD), min | 30 (16) | 39 (22) | 29 (7) | 0.27^b^ |
| Peak, mean (SD), mmol/L | 2.0 (0.6) | 1.2 (0.3)^a^ | 1.5 (0.4)^a^ | < 0.001^b^ |
| Mean (SD), mmol/L |  |  |  |  |
| Baseline | 0.2 (0.2) | 0.2 (0.3) | 0.3 (0.3) | < 0.001^c^ |
| 15 minutes | 1.6 (0.7) | 0.9 (0.3) | 1.2 (0.4) |  |
| 30 minutes | 1.9 (0.5) | 1.1 (0.3) | 1.4 (0.4) |  |
| 60 minutes | 1.4 (0.3) | 1.0 (0.4) | 1.1 (0.4) |  |
| 90 minutes | 1.0 (0.3) | 0.9 (0.3) | 0.8 (0.3) |  |
| 120 minutes | 0.5 (0.2) | 0.7 (0.3) | 0.6 (0.3) |  |
| 180 minutes | 0.5 (0.3) | 0.5 (0.3) | 0.6 (0.3) |  |
| 240 minutes | 0.6 (0.3) | 0.5 (0.3) | 0.6 (0.3) |  |

AUC, area under the curve; β-OHB, beta-hydroxybutyrate; iAUC, incremental area under the curve

^a^ Significantly different from “KE4”.
^b^ *p* value derived from linear mixed model with condition and visit order included as fixed factors, a random intercept for participant, and a planned contrast between “KE4” and each of the other supplements (“Ketone-IQ”, “Kenetik”) in case of overall statistical significance.	^c^ *p* value denotes interaction effect derived from linear mixed model with condition, time, the interaction thereof, and the visit order included as fixed factors, the baseline value as a covariate, and a random intercept for participant.

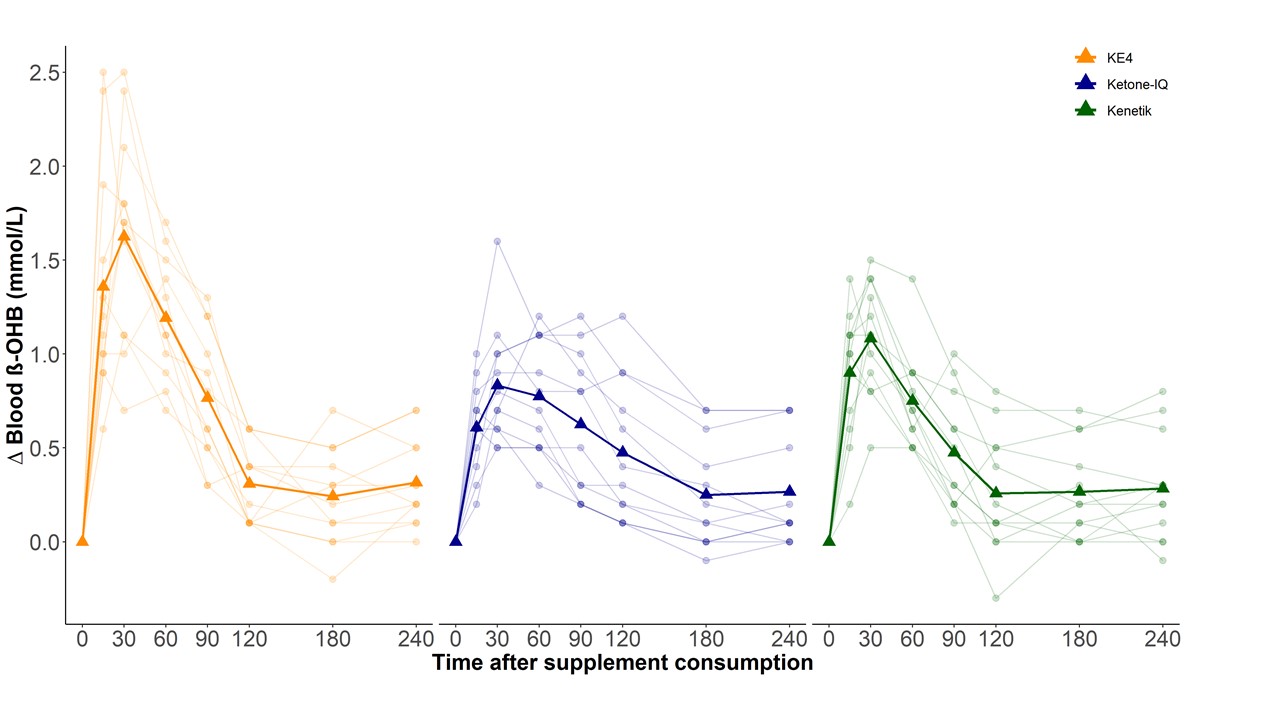

**Supplementary Figure 1**. Response of blood beta-hydroxybutyrate (β-OHB) concentration following consumption of three different ketone supplements (“KE4” left/orange; “Ketone-IQ”, center/blue; “Kenetik”, right/green) across 240 minutes. Supplements were consumed in the fasted state; β-OHB was assessed via finger prick capillary blood samples. Individual data (transparent lines and circles) alongside means (bold lines and triangles) presented (N = 12).

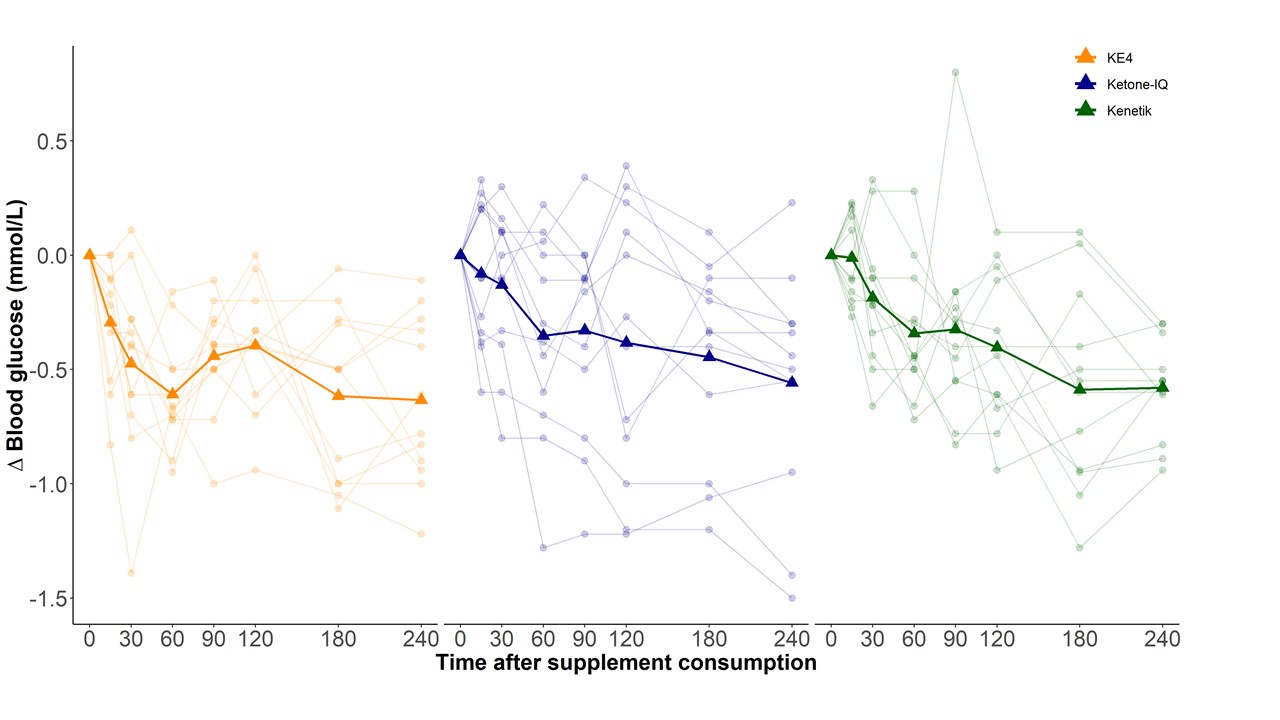

**Supplementary Figure 2**. Response of blood glucose concentration following consumption of three different ketone supplements (“KE4” left/orange; “Ketone-IQ”, center/blue; “Kenetik”, right/green) across 240 minutes. Supplements were consumed in the fasted state; glucose was assessed via finger prick capillary blood samples. Individual data (transparent lines and circles) alongside means (bold lines and triangles) presented (N = 12).

**Supplementary** **Table 4. Blood glucose response following different ketone supplements**

|  | “KE4” | “Ketone-IQ” | “Kenetik” | *p* value |
| --- | --- | --- | --- | --- |
| AUC, mean (SD), mmol/L x 240 min | 1148 (126) | 1138 (107) | 1181 (88) | 0.56^a^ |
| iAUC, mean (SD), mmol/L x 240 min | -118 (51) | -81 (93) | -94 (49) | 0.26^a^ |
| Mean (SD), mmol/L |  |  |  |  |
| Baseline | 5.3 (0.6) | 5.1 (0.4) | 5.3 (0.5) | 0.11^b,c^ |
| 15 minutes | 5.0 (0.5) | 5.0 (0.4) | 5.3 (0.5) |  |
| 30 minutes | 4.8 (0.7) | 5.0 (0.5) | 5.1 (0.5) |  |
| 60 minutes | 4.7 (0.6) | 4.7 (0.5) | 5.0 (0.5) |  |
| 90 minutes | 4.8 (0.5) | 4.8 (0.6) | 5.0 (0.4) |  |
| 120 minutes | 4.9 (0.6) | 4.7 (0.6) | 4.9 (0.3) |  |
| 180 minutes | 4.7 (0.6) | 4.6 (0.5) | 4.7 (0.5) |  |
| 240 minutes | 4.6 (0.6) | 4.5 (0.5) | 4.7 (0.5) |  |

AUC, area under the curve; iAUC, incremental area under the curve

^a^ *p* value derived from linear mixed model with condition and visit order included as fixed factors, a random intercept for participant, and a planned contrast between “KE4” and each of the other supplements (“Ketone-IQ”, “Kenetik”) in case of overall statistical significance.	^b^ *p* value denotes interaction effect derived from linear mixed model with condition, time, the interaction thereof, and the visit order included as fixed factors, the baseline value as a covariate, and a random intercept for participant.
^c^ There were statistically significant main effects of time and condition (both *p* < 0.001).

**Supplementary Table 5. Blood pressure response following different ketone supplements**

|  | “KE4” | | “Ketone-IQ” | | “Kenetik” | | *p* value^a^ | |
| --- | --- | --- | --- | --- | --- | --- | --- | --- |
|  | Systolic | Diastolic | Systolic | Diastolic | Systolic | Diastolic | Systolic | Diastolic |
| Blood pressure,  mean (SD), mmHg |  |  |  |  |  |  |  |  |
| Baseline | 112 (11) | 70 (8) | 113 (10) | 69 (12) | 113 (11) | 69 (7) | 0.88 | 0.72 |
| 15 minutes | 112 (10) | 70 (7) | 117 (13) | 70 (10) | 115 (12) | 71 (7) |  |  |
| 30 minutes | 110 (9) | 70 (10) | 114 (11) | 70 (11) | 114 (15) | 68 (9) |  |  |
| 60 minutes | 112 (9) | 70 (8) | 113 (11) | 69 (11) | 114 (16) | 73 (11) |  |  |
| 90 minutes | 113 (10) | 70 (10) | 113 (10) | 68 (13) | 116 (13) | 72 (9) |  |  |
| 120 minutes | 110 (12) | 71 (8) | 111 (15) | 68 (15) | 109 (12) | 70 (11) |  |  |
| 180 minutes | 106 (11) | 68 (10) | 113 (8) | 69 (12) | 112 (14) | 72 (9) |  |  |
| 240 minutes | 111 (9) | 69 (8) | 113 (10) | 71 (8) | 112 (15) | 71 (8) |  |  |

^a^ *p* value denotes interaction effect derived from linear mixed model with condition, time, the interaction thereof, and the visit order included as fixed factors, the baseline value as a covariate, and a random intercept for participant.

**Supplementary Table 6. Heart rate response following different ketone supplements**

|  | “KE4” | “Ketone-IQ” | “Kenetik” | *p* value^a^ |
| --- | --- | --- | --- | --- |
| Heart rate,  mean (SD), bpm |  |  |  |  |
| Baseline | 65 (13) | 64 (15) | 67 (14) | 0.04 |
| 15 minutes | 65 (13) | 64 (14) | 66 (13) |  |
| 30 minutes | 65 (12) | 64 (15) | 65 (12) |  |
| 60 minutes | 65 (14) | 64 (14) | 65 (12) |  |
| 90 minutes | 64 (13) | 62 (14) | 64 (12) |  |
| 120 minutes | 63 (13) | 63 (14) | 64 (13) |  |
| 180 minutes | 62 (13) | 62 (14) | 61 (11) |  |
| 240 minutes | 62 (13) | 63 (13) | 61 (11) |  |

^a^ *p* value denotes interaction effect derived from linear mixed model with condition, time, the interaction thereof, and the visit order included as fixed factors, the baseline value as a covariate, and a random intercept for participant.

**Supplementary Table 7. Gastrointestinal distress following different ketone supplements**

| VAS score [1 to 10], median (range) | “KE4” | “Ketone-IQ” | “Kenetik” |
| --- | --- | --- | --- |
| Nausea | 0.4 (0.0 to 3.9) | 0.0 (0.0 to 0.3) | 0.0 (0.0 to 1.1) |
| Urge to vomit | 0.0 (0.0 to 5.5) | 0.0 (0.0 to 0.2) | 0.0 (0.0 to 1.1) |
| Bloating | 0.0 (0.0 to 2.4) | 0.0 (0.0 to 1.0) | 0.0 (0.0 to 2.4) |
| Belching | 0.0 (0.0 to 5.2) | 0.0 (0.0 to 0.6) | 0.0 (0.0 to 5.2) |
| Cramping | 0.0 (0.0 to 1.6) | 0.0 (0.0 to 1.0) | 0.0 (0.0 to 2.2) |

VAS, visual analogue scale

**Supplementary Table 8. Hunger and fullness following different ketone supplements**

| VAS score [1 to 10], mean (SD) | “KE4” | “Ketone-IQ” | “Kenetik” | *p* value^a^ |
| --- | --- | --- | --- | --- |
| How hungry do you feel? | 5.8 (1.9) | 5.4 (1.9) | 5.7 (1.4) | 0.79 |
| How satisfied do you feel? | 2.7 (1.8) | 2.7 (1.6) | 2.7 (1.6) | 0.99 |
| How full do you feel? | 2.0 (1.7) | 2.2 (1.5) | 1.9 (1.5) | 0.86 |
| How much do you think you can eat? | 7.1 (1.3) | 6.8 (1.2) | 7.0 (0.9) | 0.81 |

VAS, visual analogue scale

^a^ *p* value derived from linear mixed model with condition and visit order included as fixed factors, a random intercept for participant, and a planned contrast between “KE4” and each of the other supplements (“Ketone-IQ”, “Kenetik”) in case of overall statistical significance.

**Supplementary Table 9. Analysis of blood glucose across 60 minutes following ketone supplements**

|  | “KE4” | “Ketone-IQ” | “Kenetik” | *p* value^a^ |
| --- | --- | --- | --- | --- |
| At 60 minutes, mean (SD), mmol/L | 4.7 (0.6) | 4.7 (0.5) | 5.0 (0.5) | 0.27 |
| Change from baseline, mean (SD), mmol/L | -0.6 (0.2) | -0.4 (0.4)^b^ | -0.3 (0.3)^c^ | 0.09 |
| AUC, mean (SD), mmol/L x 60 min | 292 (34) | 295 (24) | 309 (28) | 0.27 |
| iAUC, mean (SD), mmol/L x 60 min | -24 (13) | -11 (16)^d^ | -9 (11)^d^ | 0.02 |

AUC, area under the curve; iAUC, incremental area under the curve

^a^ *p* value derived from linear mixed model with condition and visit order included as fixed factors, a random intercept for participant, and a planned contrast between “KE4” and each of the other supplements (“Ketone-IQ”, “Kenetik”) in case of overall statistical significance.	^b^ Near statistical significance compared to “KE4” (*p* = 0.06).
^c^ Near statistical significance compared to “KE4” (*p* = 0.05).
^d^ Significantly different from “KE4”.

**Table 10. Predicted vs observed glucose response following ketone supplements**

|  | “KE4” | | “Ketone-IQ” | | “Kenetik” | |
| --- | --- | --- | --- | --- | --- | --- |
|  | Predicted ∆ glucose, mmol/L (95% CI) | Observed ∆ glucose, mmol/L ± SD | Predicted ∆ glucose, mmol/L (95% CI) | Observed ∆ glucose, mmol/L ± SD | Predicted ∆ glucose, mmol/L (95% CI) | Observed ∆ glucose, mmol/L ± SD |
| Average β-OHB | -0.37 (-0.44, -0.30) | -0.51  ± 0.37 | -0.32  (-0.37, -0.27) | -0.34  ± 0.32 | -0.32  (-0.37, -0.27) | -0.34  ± 0.31 |
| Peak β-OHB | -0.53 (-0.65, -0.42) |  | -0.38 (-0.44, -0.32) |  | -0.42 (-0.49, -0.34) |  |

Note: Predicted glucose values are based on the previously published meta-regression by Falkenhain et al. (6), in which the decrease (∆) in blood glucose averaged across the post-supplementation period (i.e., following ingestion of an exogenous ketone supplement in the fasted state) was predicted based on either average or peak beta-hydroxybutyrate (β-OHB) values observed across the same timeframe. The observed decrease in blood glucose is based on the supplements employed in our study and was derived similarly (i.e., averaged across the 240-minute post-supplementation period following ingestion of each ketone supplement in the fasted state).

**Supplementary Table 11. Blood beta-hydroxybutyrate and glucose following different ketone supplements separated by measurement device**

|  | “KE4” | | “Ketone-IQ” | | “Kenetik” | |
| --- | --- | --- | --- | --- | --- | --- |
| Monitor | Keto-Mojo | Abbott | Keto-Mojo | Abbott | Keto-Mojo | Abbott |
| β-OHB, mean (SD), mmol/L |  |  |  |  |  |  |
| Baseline | 0.3 (0.2) | 0.1 (0.1) | 0.3 (0.3) | 0.1 (0.0) | 0.4 (0.3) | 0.1 (0.1) |
| 15 minutes | 1.8 (0.7) | 1.2 (0.2) | 0.9 (0.3) | 0.8 (0.1) | 1.3 (0.3) | 0.9 (0.5) |
| 30 minutes | 1.9 (0.4) | 1.7 (0.8) | 1.3 (0.2) | 0.8 (0.2) | 1.5 (0.3) | 1.1 (0.6) |
| 60 minutes | 1.5 (0.2) | 1.3 (0.4) | 1.2 (0.3) | 0.7 (0.3) | 1.1 (0.3) | 0.9 (0.5) |
| 90 minutes | 1.1 (0.2) | 0.8 (0.4) | 1.1 (0.2) | 0.6 (0.3) | 0.9 (0.2) | 0.5 (0.5) |
| 120 minutes | 0.6 (0.2) | 0.4 (0.2) | 0.9 (0.2) | 0.4 (0.2) | 0.7 (0.3) | 0.3 (0.3) |
| 180 minutes | 0.6 (0.2) | 0.2 (0.1) | 0.7 (0.1) | 0.2 (0.1) | 0.7 (0.2) | 0.2 (0.2) |
| 240 minutes | 0.7 (0.2) | 0.2 (0.1) | 0.7 (0.1) | 0.2 (0.1) | 0.7 (0.2) | 0.2 (0.2) |
| Glucose, mean (SD), mmol/L |  |  |  |  |  |  |
| Baseline | 5.6 (0.2) | 4.6 (0.2) | 5.2 (0.4) | 4.9 (0.4) | 5.5 (0.2) | 4.7 (0.7) |
| 15 minutes | 5.3 (0.4) | 4.4 (0.2) | 5.1 (0.5) | 4.9 (0.4) | 5.5 (0.2) | 4.7 (0.5) |
| 30 minutes | 5.2 (0.5) | 4.1 (0.2) | 5.0 (0.4) | 4.8 (0.6) | 5.3 (0.3) | 4.6 (0.6) |
| 60 minutes | 5.0 (0.3) | 3.9 (0.2) | 4.8 (0.5) | 4.6 (0.5) | 5.1 (0.4) | 4.4 (0.7) |
| 90 minutes | 5.1 (0.3) | 4.2 (0.2) | 4.8 (0.5) | 4.6 (0.6) | 5.1 (0.3) | 4.8 (0.8) |
| 120 minutes | 5.2 (0.3) | 4.3 (0.4) | 4.8 (0.5) | 4.5 (0.6) | 5.0 (0.2) | 4.6 (0.5) |
| 180 minutes | 5.0 (0.4) | 4.1 (0.4) | 4.7 (0.5) | 4.6 (0.6) | 4.8 (0.4) | 4.4 (0.8) |
| 240 minutes | 5.0 (0.4) | 4.0 (0.3) | 4.6 (0.4) | 4.4 (0.7) | 4.7 (0.2) | 4.3 (0.8) |

β-OHB, beta-hydroxybutyrate

Note: These data separated by measurement device were not compared within conditions and participants; therefore, direct comparisons do not reflect device differences.
